# Does reactogenicity after a second injection of the BNT162b2 vaccine predict spike IgG antibody levels in healthy Japanese subjects?

**DOI:** 10.1101/2021.06.08.21258444

**Authors:** Masaaki Takeuchi, Yukie Higa, Akina Esaki, Yosuke Nabeshima, Akemi Nakazono

## Abstract

Adverse reactions are more common after the second injection of messenger RNA vaccines such as Pfizer/BioNTech’s BNT162b2. We hypothesized that the degree and severity of reactogenicity after the second injection reflects the magnitude of antibody production against the SARS CoV-2 virus spike protein (spike IgG). Blood samples were obtained from 67 healthy Japanese healthcare workers three weeks after the first injection and two weeks after the second injection of the BNT162b2 vaccine to measure spike IgG levels. Using questionnaires, we calculated an adverse event (AE) score (0-11) for each participant. The geometric mean of spike IgG titers increased from 1,047 antibody units (AU/mL) (95% CI: 855−1282 AU/mL) after the first injection to 17,378 AU/mL (14,622−20,663 AU/mL) after the second injection. The median AE score increased from 2 to 5. Spike IgG levels after the second injection were negatively correlated with age and positively correlated with spike IgG after the first injection. AE scores after the second injection were not significantly associated with log-transformed spike IgG after the second injection, when adjusted for age, sex, and log-transformed spike IgG after the first injection. Although the sample size was relatively small, reactogenicity after the second injection may not accurately reflect antibody production.

## Introduction

Effective and appropriate coronavirus disease 2019 (Covid-19) vaccination is the most promising strategy for controlling the spread of SARS-CoV2 infection on a worldwide basis.^1-3^ Currently, nucleoside-modified messenger RNA (mRNA) vaccines encoding SARS-CoV2 full-length spike and adenoviral vector vaccines have been used mainly in Western countries, and vaccination levels in the United States, England, and Israel are now reaching 45% - 63%, dramatically reducing the infection rate.^4-7^ In Japan, the number of Covid-19 infections is increasing, but the vaccination rate is still very low (<3%) due to a limited supply of vaccines.

Local and systemic reactogenicity after the second injection of mRNA vaccines is more common than after the first injection.^1,2,8^ The second injection has a booster effect that produces a substantial antibody titers against SARS-CoV2 spike antigen. Thus, we hypothesized that the degree of reactogenicity following the second injection could be an indicator of the level of SARS-CoV2 spike antibody production. Accordingly, the aim of this study was to investigate whether local and systemic reactogenicity after the second injection of an mRNA vaccine reflects subsequent SARS-CoV2 spike antibody levels in healthy recipients.

## Materials and Methods

### Study participants

This was a prospective, longitudinal, observational study in a single center. The study was approved by the institutional review board of the University of Occupational and Environmental Health, School of Medicine (approval number: UOEHCRB21-023). Currently, the only vaccine available in Japan is the BNT162b2 mRNA Covid-19 vaccine (Pfizer/BioNTech). The Japanese government started to distribute this vaccine to healthcare workers in February 2021, and our university hospital received the vaccine in the middle of March 2021. The hospital chairman decided to administer the first dose of BNT162b2 mRNA Covid-19 vaccine (30 μg per dose injected into the deltoid muscle) to hospital employees during the fourth week of March and the second dose during the third week of April (April 12 to April 16). Since we received the ethical approval for the study on April 12 the time available to acquire informed consent and to conduct the study was quite short, so we asked mainly healthcare workers in our hospital department to participate in the study. Written informed consent was obtained from all 69 participants.

### Antibody test

Blood samples were obtained from participants before the second dose of the BNT162b2 mRNA vaccination (median: 20 days [interquartile range (IQR): 20 to 21 days] after the first dose) and 2 weeks after the second dose (median: 13 days [IQR: 11 to 14 days] after the second dose). A SARS-CoV2 IgG assay was performed with chemiluminescent immunoanalysis of microparticles used for quantitative detection of IgG antibodies against the spike protein of the SARS CoV-2 virus (spike IgG) employing the Architect system (Abbott Diagnostics) with a cut-off <50 antibody units/mL (AU/mL) in both blood samples. To exclude the possibility of previous Covid-19 infection, we also measured IgG antibodies against the nucleocapsid protein of Covid-19 (Abbott Diagnostics) using the second blood sample. The negative cut-off value for anti-nucleocapsid protein IgG was < 1.4 AU/mL.

### Reactogenicity

All participants were asked to respond to questionnaires after both the first and second blood samples regarding local and systemic adverse effects to the vaccine injections. Questionnaires included a previous history of allergic reactions, severe adverse effects such as anaphylactic reactions, local reactogenicity such as pain at the injection site, swelling, lymph node swelling, and systemic reactogenicity, including fatigue, headache, myalgia, arthralgia, chills, and fever (body temperature > 38 °C). To estimate reactogenicity, we calculated an adverse effect score (AE score). When local and systemic reactogenicity, except for fever, was present, we scored it as 1. Regarding fever, a score of 1 was given when body temperature increased to ≥38.0 °C but <38.4°C. A fever score of 2 corresponded to a body temperature from ≥38.4 °C to <38.9°C. Development of a body temperature ≥38.9°C was scored as 3.^2,3^ We summed all scores and calculated AE scores after the first and second injection (range: 0 to 11).

The primary outcome was to determine whether the AE score after the second injection was associated with spike IgG levels two weeks after the second injection.

### Statistical analysis

Levels of spike IgG were expressed as the median, IQR, and the geometric mean and 95% confidence intervals calculated using the Student’s t test distribution on log-transformed data. AE score was shown as the median and IQR. Categorical data were presented by number and percentage. Given the small number of participants, we did not perform formal statistical comparisons between groups. Spearman’s correlation analysis was used to compare spike IgG levels after the first and second injections and between spike IgG after the second injection and age. Correlation analysis was further performed in subgroups stratified by gender. To predict the level of log-transformed spike IgG two weeks after the second injection, multivariate linear regression analysis was performed, using age, sex, log-transformed spike IgG three weeks after the first injection, and AE score after the second injection as covariates. A two-sided p-value < 0.05 was considered significant. All statistical analyses were conducted using R software version 4.0.4 (R foundation for Statistical Computing, Vienna).

## Results

Two participants did not provide a second blood sample within the specified time window; thus, we excluded them from the analysis. The final study population comprised 67 participants (median age: 43 years, IQR: 32 to 51 years, 21 men). Six subjects reported previous histories of allergic reactions.

### Reactogenicity

No one developed a severe adverse reaction after the vaccinations. Figure 1A presents local and systemic reactogenicity after the first and second injections. Except for myalgia, the prevalence of all other reactogenicities increased from the first injection to the second injection, resulting in a higher AE score after the second injection (median: 5, IQR: 2−6) than after the first injection (median: 2, IQR: 1−3). Since previous studies showed higher reactogenicities in younger subjects ^2,9^ we divided subjects into two groups stratified by age and compared reactogenicities after the second injection. AE scores were higher in subjects who were < 50 years (median: 5, IQR: 4−6) compared with subjects ≥ 50 years (median: 2, IQR: 1−3).

**Figure 1:**
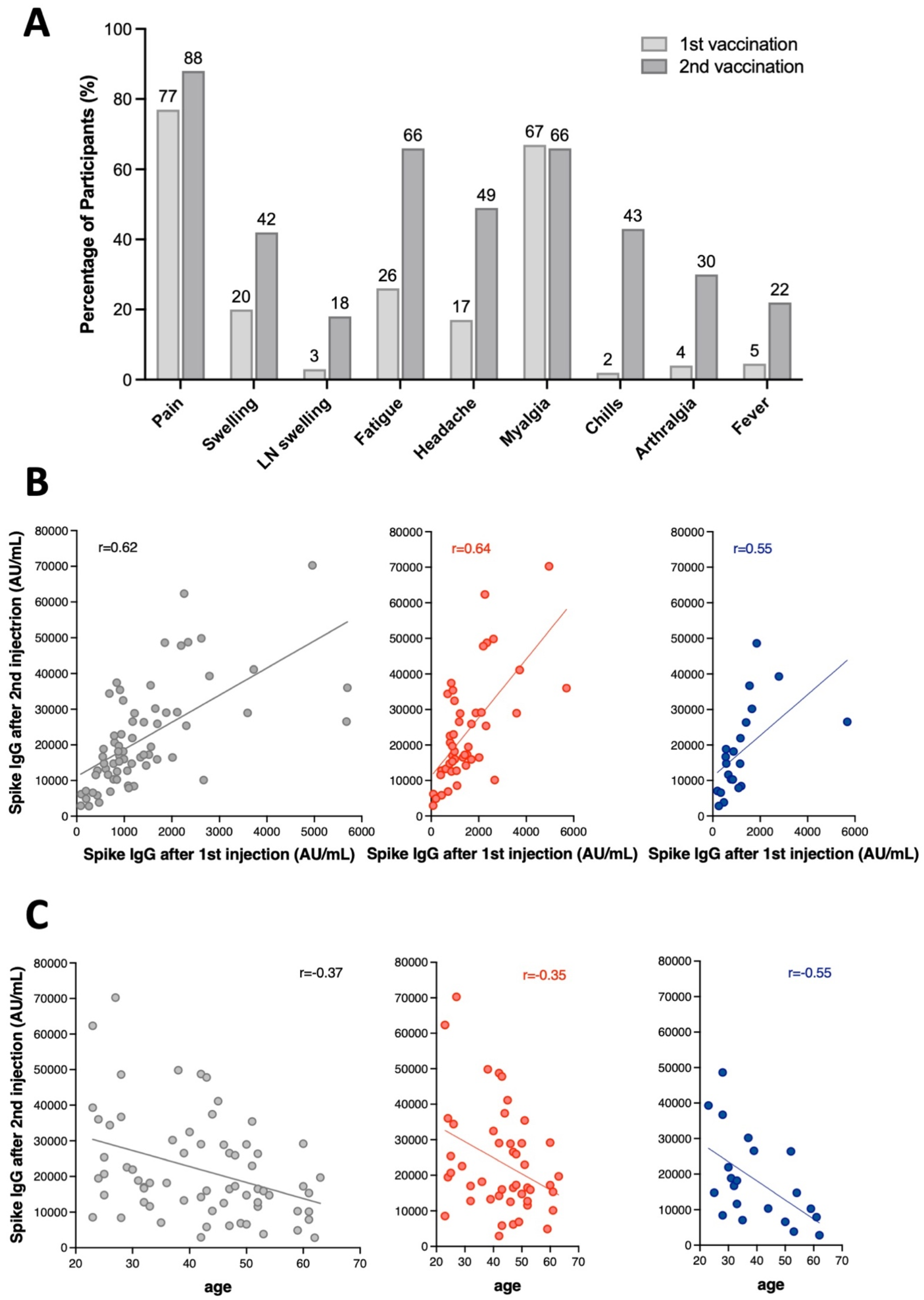
A, Local and systemic reactions after the first and second injections of the Pfizer/BioNTech BNT162b2 vaccine among all subjects. Fever was defined as a body temperature > 38 °C after a vaccine injection. B, A linear correlation between spike IgG level after the first injection and that after the second injection in all subjects (left), in female subjects (middle), and in male subjects (right). C, A linear correlation between age and spike IgG level after the second injection in all subjects (left), in female subjects (middle), and in male subjects (right).

### Antibodies

None of the subjects were positive for anti-nucleocapsid IgG (median: 0.04 AU/mL, IQR: 0.02−0.07 AU/mL). All subjects were positive for spike IgG (> 50 AU/mL), for which the median, IQR, geometric mean and its 95% confidence interval after the first injection were 1065 AU/mL, 741 −1693 AU/mL, 1047 AU/mL, and 855−1282 AU/mL, respectively. After the second injection, corresponding values increased to 17259 AU/mL, 12088 −28999 AU/mL, 17378 AU/mL, and 14622−20663 AU/mL, respectively, a 16-fold increase of spike IgG from the first injection to the second. There was a positive correlation between first spike IgG levels and second spike IgG levels (Figure 1B). There was also a negative correlation between second spike IgG levels and age (Figure 1C).

### Multivariate linear regression analysis

Multivariate linear regression analysis revealed that age and log-transformed spike IgG after the first injection were significantly associated with log-transformed spike IgG after the second injection. However, AE score after the second injection and sex were not significantly associated with log-transformed spike IgG after the second injection (Table 1). After removing those two variables from the model, the final model yielded an F-statistic of 52.4, residual standard error of 0.29, and an adjusted R^2^ of 0.61.

**Table 1:** Multivariate linear regression analysis of the association of log-transformed spike IgG two weeks after the second injection of the Pfizer/BioNTech BNT162b2 mRNA vaccine.

| **variables** | **Beta coefficient** | **Standard Error** | **t-value** | **p-value** |
| --- | --- | --- | --- | --- |
| age | -0.0057 | 0.0021 | -2.677 | 0.009 |
| male gender | -0.0939 | 0.0501 | -1.875 | 0.066 |
| log spike IgG after the first injection | 0.5496 | 0.0668 | 8.096 | <0.001 |
| AE score after the second injection | 0.0174 | 0.0108 | 1.622 | 0.110 |

Residual standard error: 0.1864
Adjusted R<sup>2</sup>: 0.6345
F-statistic: 29.64, p<0.001

## Discussion

The main findings of this study can be summarized as follows: (1) the prevalence of local and systemic reactogenicity increased approximately 2.5 times after the second injection, compared to the first injection, and its severity was more significant in younger people; (2) spike IgG titers became positive in all participants after the first injection, and those titers increased 16-fold after the second injection; (3) spike IgG titers after the second injection were negatively correlated with age, and positively correlated with spike IgG after the first injection; (4) multivariate linear regression analysis revealed that the degree of reactogenicity after the second injection was not a significant predictor of spike IgG levels after the second injection after adjusting for age, gender, and spike IgG levels after the first injection.

None of the subjects in this study showed positive results for nucleocapsid IgG; thus, we believe that all participants were Covid-19 naïve subjects.^10^ Although the degree of adverse effects after the first injection was similar to those in a previous publication from western countries,^2,3,11^ the prevalence of reactogenicity after the second injection was higher. Although proposed causes may be related to racial differences and geographic differences of non COVID-19 corona virus exposure, we should consider a fixed dosing regimen as a potential cause of different prevalence of adverse reactions. Dose-dependent increases in adverse effects have been observed with mRNA vaccines ^1-3^ and the current recommended dose of BNT162b2 mRNA vaccine (30 μg), which was determined for western subjects, would result in a higher dose for people with smaller body mass, such as Japanese.

The geometric mean of spike IgG after the second injection was 17378 AU/mL, which was higher than corresponding values measured in a naïve Spanish COVID-19 population, ^12^ but similar to another study. ^13^ In addition, the range of values was quite broad, from 2826 AU/mL to 70272 AU/mL. Although there is no definite cut-off of value for spike IgG sufficient to prevent COVID-19 infection, the FDA authorized only the use of high-titer COVID-19 convalescent plasma (spike IgG > 840 AU/mL), for treatment of hospitalized patients with COVID-19 infection, early in the course of the pandemic.^14^ Prendecki et al. reported that one COVID-19 naïve individual whose spike IgG titer was 61.8 AU/mL, developed symptomatic COVID-19 infection 5 weeks after the first dose of BNT162b2 vaccine.^15^ The authors also stated that spike IgG titers < 250 AU/mL might not be sufficient for virus neutralization. The lowest IgG titers observed in this study exceeded the latter value by nearly 10-fold. Spike IgG levels after the second injection were higher in subjects < 50 years than in those ≥ 50 years. Previous studies did not show age-dependent efficacy of BNT162b2 for subsequent COVID-19 infection.^3,6^ Thus, these results raise concern regarding the recommended dose of BNT162b2 mRNA vaccine in younger Japanese people. The current infection situation in Japan makes dose response trials with BNT162b2 vaccine almost impossible. However, if the level of spike IgG three weeks after the first vaccination is > 1000 AU/mL, the second injection dose could be reduced in younger people, which would enhance the Japanese vaccination rate.

Although efficacy and adverse effects of vaccines appear opposite, they may be related via the strength of the immune response to the vaccine. However, there was no significant association between AE score after the second injection and spike IgG levels after adjusting for age, gender, and spike IgG level after the first injection. Although the relatively small sample size could produce false negative results, adverse effects after the second injection might be dose-dependent immune reactions after repeated exposure to excipients to stabilize lipid nanoparticles, such as PEG2000, which serves as the vehicle for the vaccine mRNA.^8^ Age was negatively correlated with spike IgG production, which agrees with previous studies.^3,9^ Spike IgG levels after the first injection were also positively correlated with spike IgG production after the second injection. However, the model including age and spike IgG after the first vaccination was not sufficiently robust to predict the amount of spike IgG after the second injection with confidence.

Small sample size limited the generalizability of our results. Since our results were derived from healthy Japanese healthcare workers, further studies should examine subjects with comorbidities and more aged populations.^16,17^ A longitudinal study to determine spike IgG levels after vaccination is another important topic for intensive research.

In summary, reactogenicity after the second injection of BNT162b2 mRNA vaccine was common, especially in young to middle aged Japanese healthcare workers. However, its intensity was not closely correlated with subsequent spike IgG levels, after adjusting for age, sex, and spike IgG levels after the first injection. Thus, reactogenicity after the second injection is probably not a reliable measure of antibody production.

## Data Availability

Data will be available with a reasonable request.

## Conflict of interest

MT received a research grant from Abbott. Other authors declare that they have no conflicts of interest.

## Data availability statement

Data supporting the findings of this study are available from the corresponding author upon request.

## Notes

### Author Declarations

The study was approved by the institutional review board of the University of Occupational and Environmental Health, School of Medicine (approval number: UOEHCRB21-023).

